# The impact of same-day antiretroviral therapy initiation under the WHO Treat-All policy

**DOI:** 10.1101/2020.05.04.20090068

**Authors:** Bernhard Kerschberger, Andrew Boulle, Rudo Kuwengwa, Iza Ciglenecki, Michael Schomaker

## Abstract

**Background:** Rapid initiation of antiretroviral therapy (ART) is recommended for people living with HIV, with the option to start treatment on the day of diagnosis (same-day-ART). However, the effect of same-day-ART remains unknown in realistic public sector settings.

**Methods:** We established a cohort of ≥16-year-old patients who initiated first-line ART under Treat-All in Eswatini between 2014-2016, either on the day of HIV care enrolment (same-day-ART) or 1–14 days thereafter (early-ART). Directed acyclic graphs, flexible parametric survival analysis and targeted maximum likelihood estimation (TMLE) were used to estimate the effect of same-day-ART initiation on the composite unfavourable treatment outcome (loss to follow-up;death;viral failure).

**Results:** Of 1328 patients, 839 (63.2%) initiated same-day ART. The adjusted hazard ratio of the unfavourable outcome was increased by 1.48 (95% CI:1.16–1.89) for same-day-ART compared with early-ART. TMLE suggested that after 1 year, 28.9% of patients would experience the unfavourable outcome under same-day-ART compared with 21.2% under early-ART (difference: 7.7%; 1.3–14.1%). This effect was driven by loss to follow-up and varied over time, with a higher hazard during the first year after HIV care enrolment and a similar hazard thereafter.

**Conclusions:** We found an increased risk with same-day-ART. A limitation was possible silent transfers that were not captured.

## BACKGROUND

The World Health Organization (WHO) policy for Treat-All recommends lifelong antiretroviral therapy (ART) for all people living with HIV (PLHIV) at the time of diagnosis, irrespective of immunological criteria.^1^ Despite high uptake of this policy in Africa,^2^ of 20.6 million PLHIV in Eastern and Southern Africa, treatment coverage (67%) and viral suppression (58%) remained below the UNAIDS targets in 2018, with an additional 3.0 million PLHIV needing to access treatment and achieve viral suppression.^3,4^

Accelerated ART initiation has been proposed to overcome some of these gaps.^5,6^ A systematic review found that ART initiation on the same day as HIV diagnosis or the day of treatment eligibility improved treatment uptake, HIV care retention and viral suppression.^7^ Based on this evidence, WHO released guidelines in 2017 recommending ART initiation within 7 days of HIV diagnosis (rapid ART), with the possibility of initiating treatment on the same day as HIV diagnosis (same-day ART) for patients ready to start.^8^

As HIV programmes allow for accelerated ART initiation under Treat-All and most treatment initiations already occur quickly, within 14–30 days after HIV diagnosis or care enrolment,^9–13^ the question increasingly shifts to how much faster ART can be initiated in routine resource-limited settings (RLS). This question has also been raised recently in public HIV treatment programmes in high-income countries.^14^ Concerns were specifically raised about the feasibility of same-day ART initiation in realistic public sector settings because of lack of real-world evidence and practical limitations. Firstly, evidence of the benefits of accelerated ART mainly originated from randomized trials.^7^ These trials often applied additional procedures not routinely available in RLS (e.g. accelerated counselling protocols, treatment readiness survey), used treatment eligibility criteria in use before Treat-All, restricted ART interventions to specific patient groups (e.g. non-pregnant adults) or few facilities, or applied different definitions of same-day ART.^7,15–17^ In contrast, benefits of same-day ART initiation remained uncertain in observational studies.^7^ Secondly, real-world effectiveness may be compromised because of pre-existing constraints in the public sector, such as resource limitation (e.g. human resources), overburdened health facilities and suboptimal quality of care.compromised because of pre-existing constraints in the public sector, such as resource limitation (e.g. human resources), overburdened health facilities and suboptimal quality of care.compromised because of pre-existing constraints in the public sector, such as resource limitation (e.g. human resources), overburdened health facilities and suboptimal quality of care.^18-21^

Treat-All has been implemented in a public sector setting in southern Eswatini (formerly Swaziland) since 2014, with same-day ART initiation increasingly practised.^12^ Therefore, this setting provides a unique opportunity to better understand how much faster ART should be started in a context where it is already started quickly. We aimed to answer the following questions: 1) how is same-day ART being implemented in a public sector programme applying the Treat-All approach, and 2) what is the effect of same-day ART initiation compared with early ART initiation (1–14 days after HIV care enrolment) on treatment outcomes for patients starting treatment quickly.

## METHODS

### Setting

Details of the study setting were described elsewhere.^12,22^ In brief, Eswatini has an HIV prevalence of 32% among adults aged 18–49 years, and annual TB incidence was 308 cases per 100,000 population with 75% HIV co-infection in 2017.^23,24^ Treat-All was piloted in eight primary and one secondary care public sector facilities in the predominantly rural Nhlangano health zone of the Shiselweni region. Other facilities of the region were excluded from this study as they applied the CD4 350 and 500 cells/mm^3^ treatment eligibility thresholds as recommended by national treatment guidelines.^12^ ART initiation was possible in the absence of baseline CD4 cell counts and biochemistry results.^25^ ART initiation on the day of facility-based HIV care enrolment was policy for pregnant/lactating women and encouraged for other patients in the absence of (presumptive) opportunistic infections.^25,26^ Without specific standard operating procedures in place for same-day ART initiation under Treat-All at that time, the clinician decided on the timing of ART initiation after clinical and psychological readiness assessment, the patient’s perceived readiness as well as other clinical considerations. As HIV care registration and ART initiation were performed by facility-based clinicians, same-day ART initiation (on the day of HIV diagnosis) was in practice infeasible for HIV-positive patients transferred in from non-HIV care facilities and community HIV testing sites. Led by lay counsellors, one group-counselling session and at least one individual-counselling session were recommended, and both could happen on the same day as HIV diagnosis, care enrolment and ART initiation. Adherence counselling support continued thereafter as per patients’ needs. Routine follow-up visits were scheduled at 2, 4 and 12 weeks after ART initiation and 3-monthly thereafter. Routine viral load monitoring was performed 6 and 12 months after ART initiation and annually thereafter. Patients with viral loads >1000 copies/mL received enhanced adherence counselling over 3 months and were switched to second-line ART in case of viral failure (two consecutive viral load measurements >1000 copies/mL).^27^ Patients who missed their clinical appointment for medication refills received phonic defaulter tracing with the possibility of home visits.

### Study design

This is a nested, retrospectively established cohort of adults ≥16 years old initiating standard first-line ART under the Treat-All programmatic approach in Nhlangano health zone either on the day of facility-based HIV care enrolment (same-day ART) or 1–14 days after HIV care enrolment (early ART), between 10 October 2014 and 31 March 2016. A standard first-line treatment regimen contained lamivudine, and tenofovir or zidovudine, and efavirenz or nevirapine. A patient was considered enrolled into HIV care and initiated on ART if a paper and/or electronic patient record was created. In this setting, we considered early ART as a relevant comparison group to same-day ART because this was the national policy at the time of the study.

### Analyses and main definitions

Analyses were performed with Stata 14.1 and R. Firstly, baseline characteristics were described with frequency statistics and proportions. The Pearson’s chi-squared and Mann-Whitney U test were used to compare differences in categorical and continues variables. We used multiple imputation by chained equations^28^ to deal with missing values of the measured pre-treatment variables (see Supplementary Table 1).

Secondly, we assessed predictors of same-day ART initiation compared with early ART by using multivariable Poisson regression models including all variables measured before treatment initiation (listed in Table 1 and Figure 1).

**Table 1:**
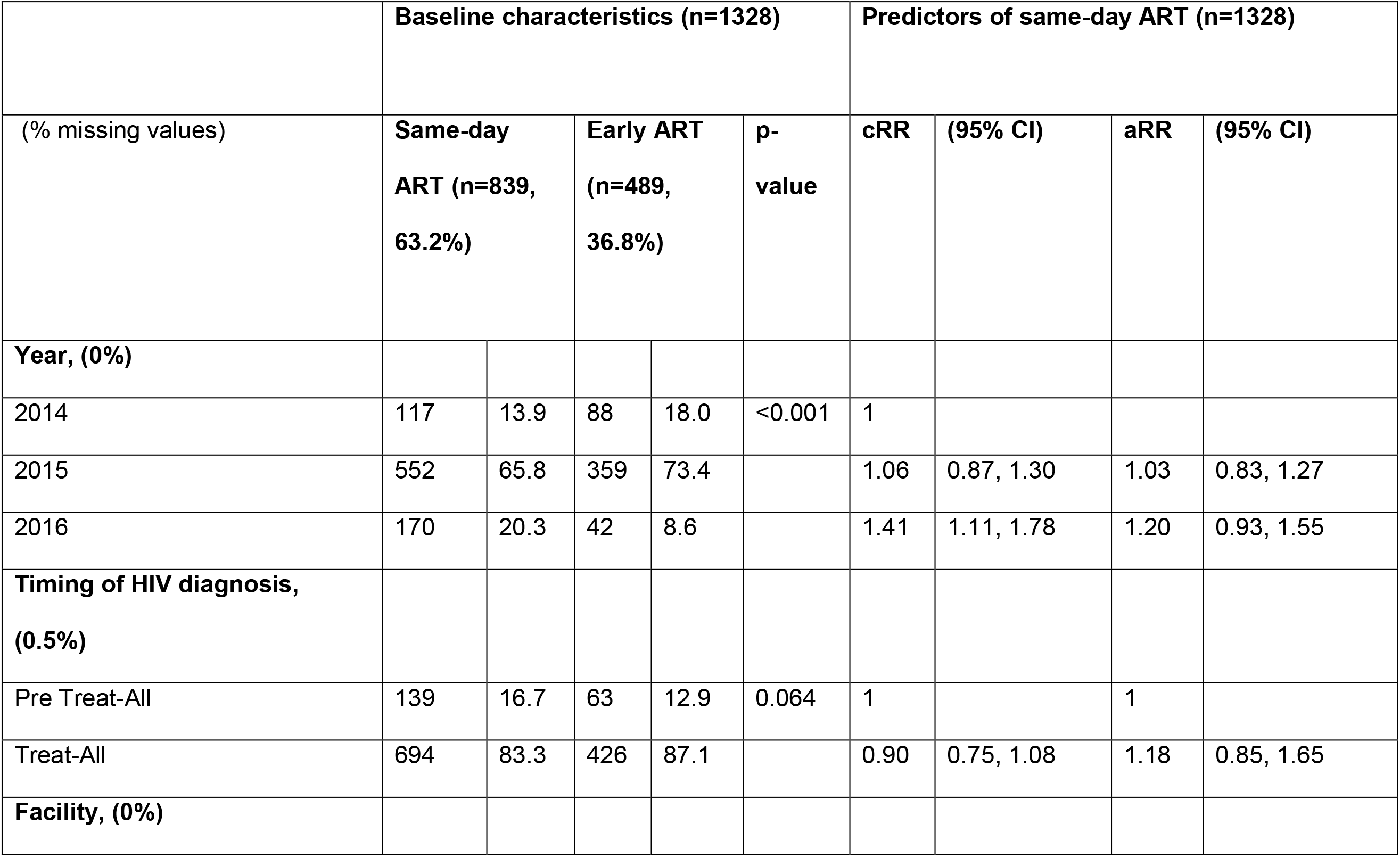
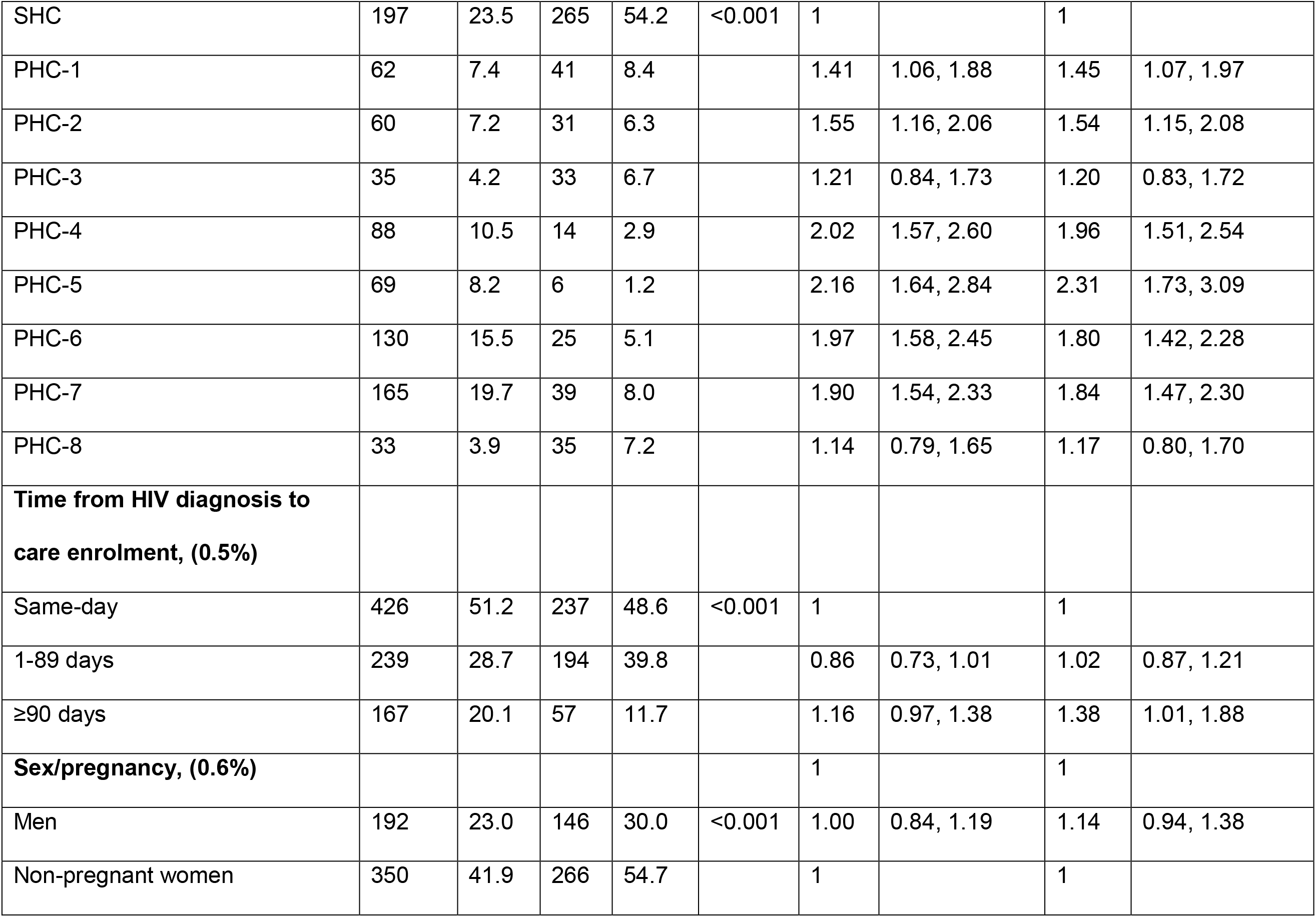
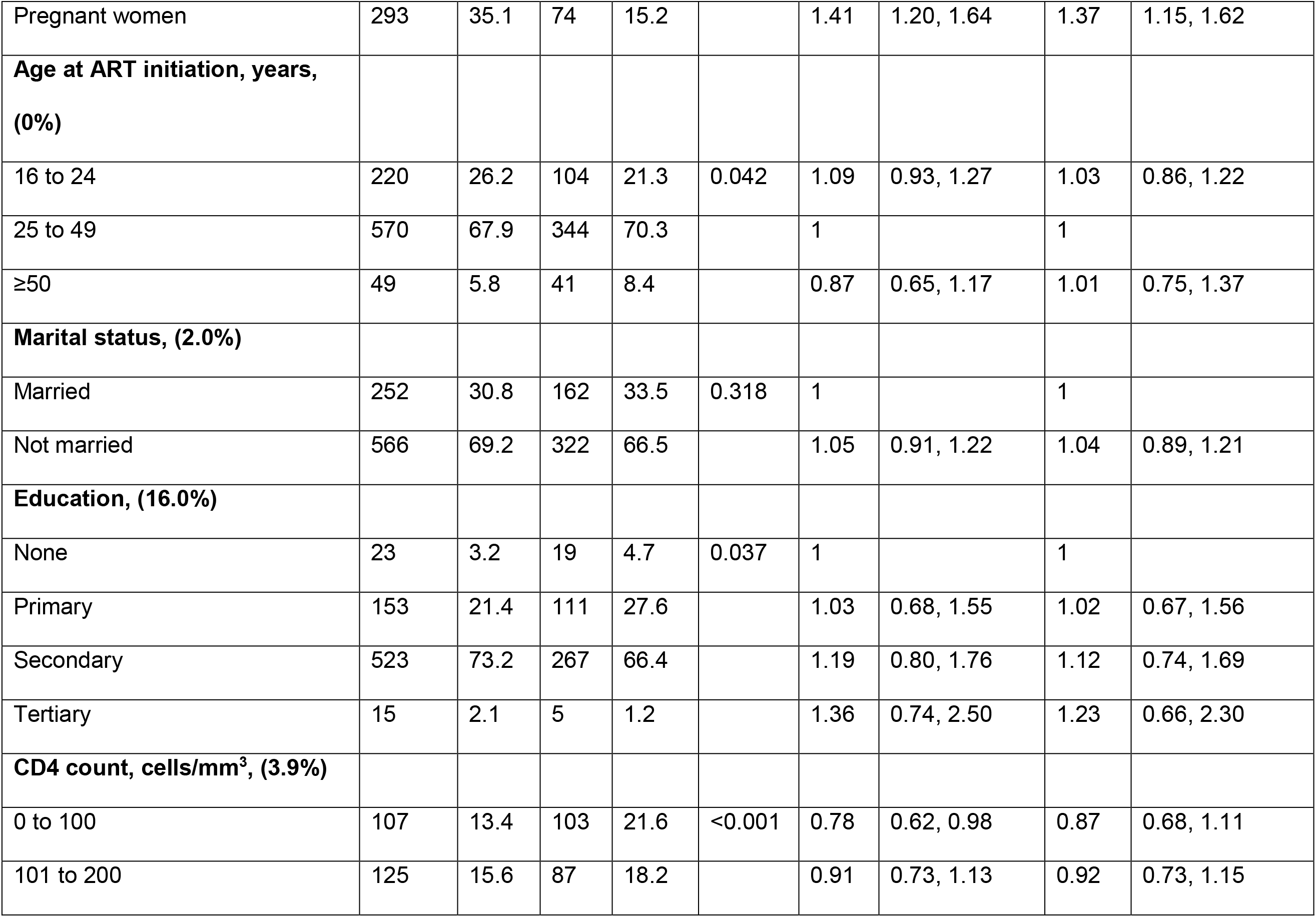
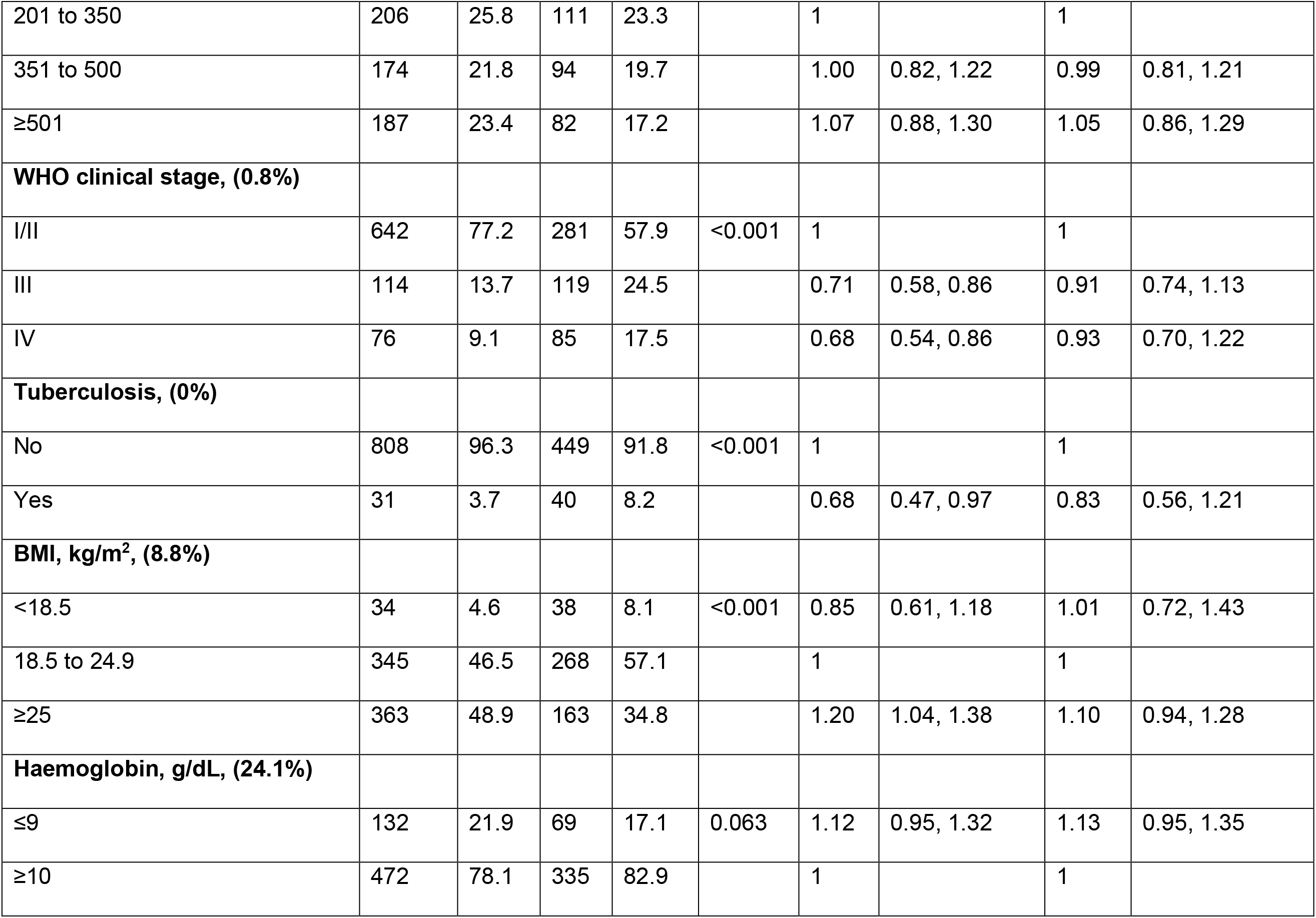
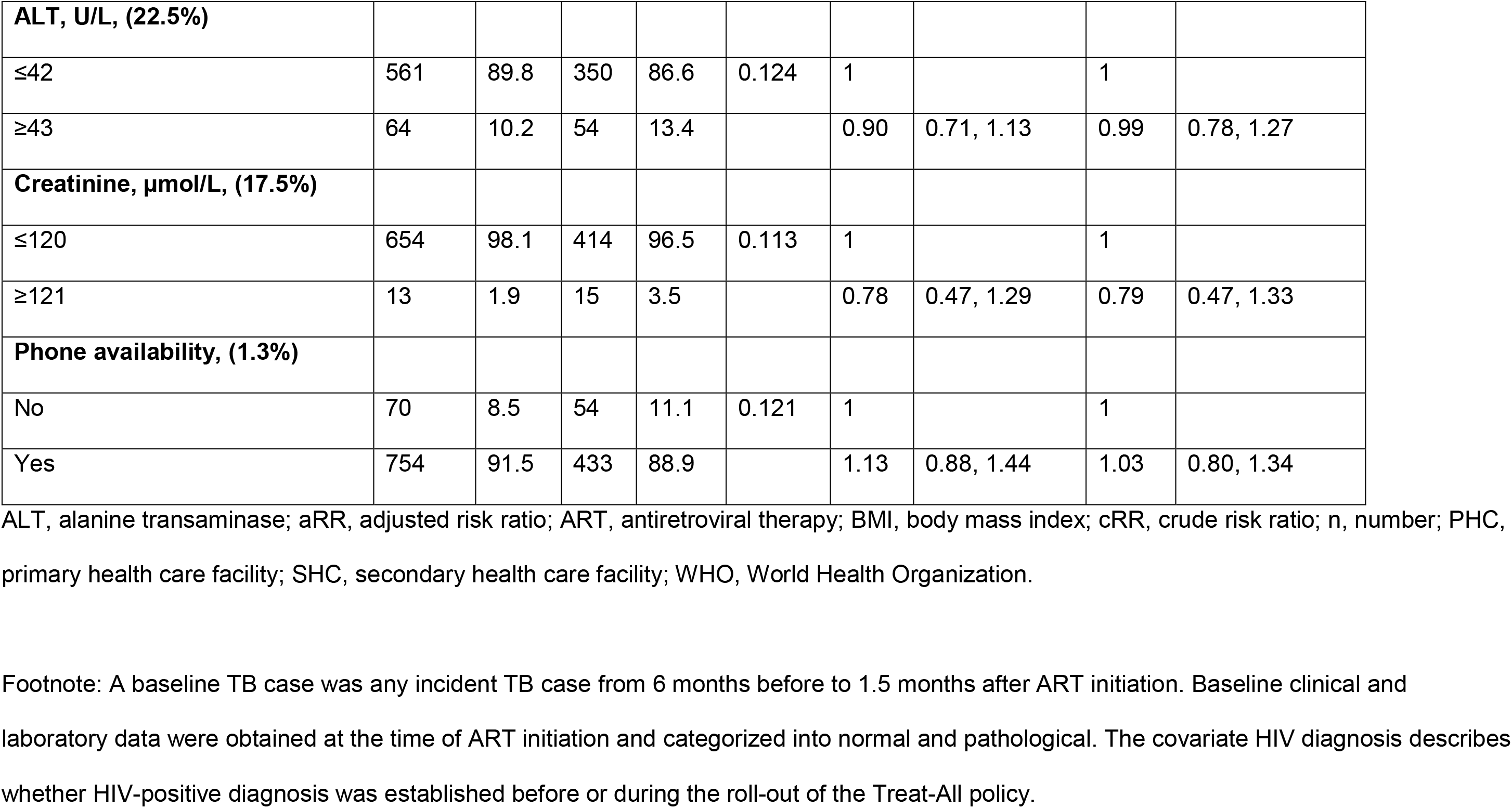
Baseline characteristics of patients initiated on ART under same-day and early ART, and predictors of same-day ART initiation, Same-day antiretroviral therapy under Treat-All, 2014–2016.

**Figure 1:**
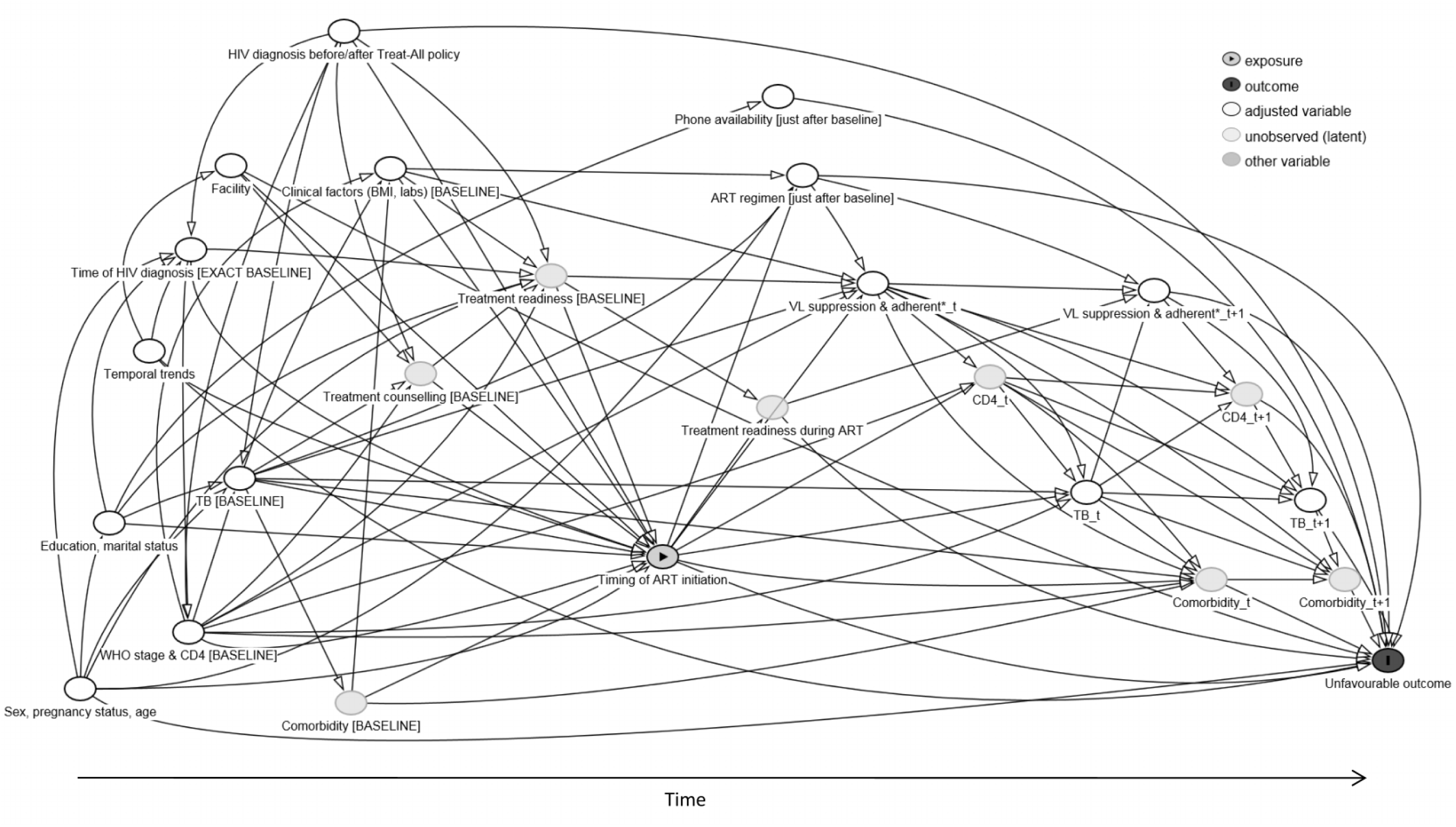
Directed acyclic graph (DAG) showing structural assumptions about the data-generating process, Same-day antiretroviral therapy under Treat-All, 2014–2016.

Thirdly, we emulated a Target Trial^29–31^ of HIV-infected patients aged ≥16 years already initiated on ART within 14 days of facility-based HIV care enrolment to estimate the causal effect^32^ of same-day ART (vs early ART) on the composite unfavourable treatment outcome of death, loss to follow-up (LTFU), viral failure and treatment switching to a second-line ART in the absence of documented viral failure. Time zero was the date of ART initiation because some captured outcomes (viral failure, treatment switch) could only have happened after ART initiation and the outcomes of death and LTFU before ART initiation were not well defined (e.g. pre-treatment visits were not recorded after care enrolment, which may lead to possible misclassification of deaths as LTFU). Therefore, our target population excluded patients starting treatment >14 days after care enrolment and patients never starting treatment for any reason (including deaths within 14 days of care enrolment).

Viral failure was defined as two consecutive viral load measurements >1000 copies/mL measured at least 5 months after ART initiation and 1.5 months apart. The composite endpoint was chosen to reflect the goals of Treat-All and the UNAIDS 90-90-90 cascade targets of keeping patients on effective (virally suppressed) ART and reduce transmission of HIV. Minimum follow-up time before database closure was 7 months. Patients were censored at the last clinic visit date when a transfer out (TFO) was recorded by the clinician and at database closure (31 October 2017). LTFU was defined as no-show to the facility for ≥6 months measured at the last clinic visit. Lacking local evidence, no assumptions were made about possible reasons of LTFU such as undocumented deaths, silent TFO, unstructured treatment interruptions and actual disengagement from care.^35–35^

We summarized our assumptions about the data-generating process in a directed acyclic graph (DAG, Figure 1); see Supplementary Technical Appendix 1 for a detailed explanation. Briefly, treatment assignment is based on various factors, including pregnancy, clinician’s preference in each facility, temporal trends, the patient’s perceived readiness and the impact of counselling, and clinical assessment including CD4 count and co-morbidities. Timing of treatment initiation may affect the composite outcome in different ways: firstly, biologically, if treatment delay would affect viral suppression and thus the development of co-morbidities and negative outcomes; secondly, earlier treatment may have a psychological impact on patients. If they do not feel ready for ART and are possibly coerced into treatment, adherence to therapy may be suboptimal and treatment may be interrupted. The DAG shows that inclusion of all visualized pre-treatment variables, and exclusion of all post-treatment variables (e.g. suppression during follow-up, ART regimen), is sufficient to identify the desired total causal effect (because all back-door paths are blocked and no mediators are being conditioned on).^36^ However, as treatment readiness and counselling, as well as some baseline co-morbidities (e.g. cryptococcal meningitis), are unmeasured, some remaining unmeasured confounding may persist in our analysis.

Based on the above assumptions, we included all measured pre-treatment variables in an adjusted flexible parametric survival analysis (Royston-Parmar models)^37,38^ to estimate the effect of same-day ART initiation on the hazard of the unfavourable outcome. We visualized the results of this model using averaged failure and hazard difference curves to compare the time to the composite unfavourable outcome between same-day and early ART.time to the composite unfavourable outcome between same-day and early ART.time to the composite unfavourable outcome between same-day and early ART.

Then, we used targeted maximum likelihood estimation (TMLE)^39,40^ to estimate the probability of experiencing the unfavourable outcome 12 months, 18 months and 24 months after ART initiation under same-day and early ART, and under no censoring, using all measured pre-treatment variables. TMLE requires estimation of the expected outcome, treatment assignment and censoring processes, given the measured covariates. We facilitated this step using extensive super learning to avoid model mis-specification (see Supplementary Table 2).^41,42^

Several supplementary analyses were performed. We compared same-day ART with rapid ART initiation defined as ART initiation 1–7 days after HIV care enrolment (rather than early ART) as per WHO recommendations. Then, the composite unfavourable outcome was decomposed to all-cause attrition (death and LTFU combined). Finally, time zero was defined as the date of HIV care enrolment (instead of ART initiation).

### Ethics

This retrospective analysis was nested within a prospective cohort study assessing the feasibility of Treat-All^12^ and was approved by the MSF ethics review board, the Eswatini National Health Research Review Board and the Human Research Ethics Committee of the University of Cape Town.

## RESULTS

Figure 2 shows the study flow chart. Of 1899 patients initiating ART, 1341 (70.6%) started treatment within 14 days after facility-based HIV care enrolment. Thirteen (1.0%) patients were removed from the analysis, as study eligibility remained unclear. Of 1328 patients remaining, 839 (63.2%) started ART on the same day as HIV care enrolment.

**Figure 2:**
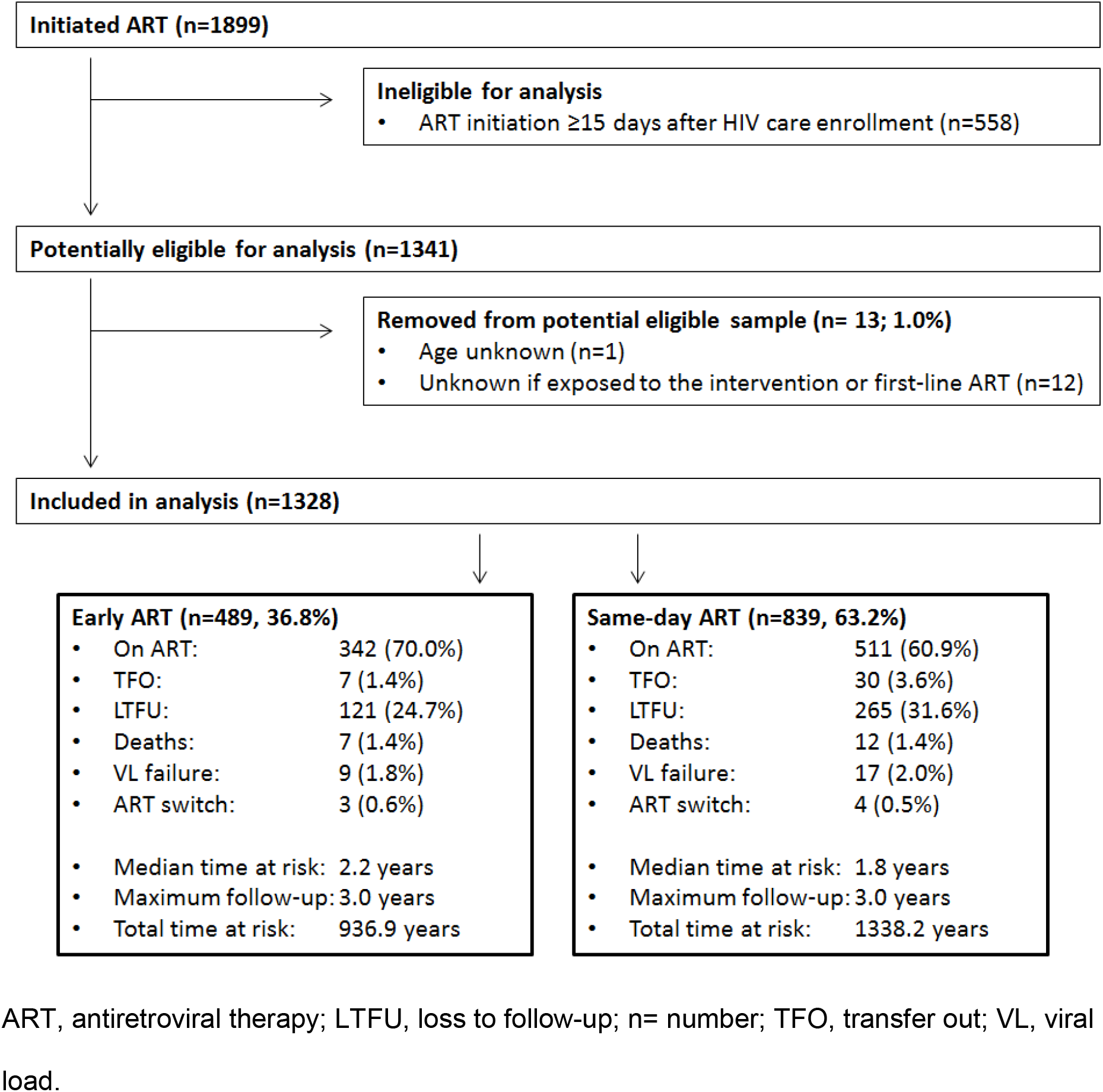
Study flow chart, Same-day antiretroviral therapy under Treat-All, 2014–2016.

### Predictors of same-day ART initiation

Table 1 shows baseline characteristics of patients starting ART same-day and early. In multivariable analysis (Table 1), the risk of same-day ART initiation was higher for 6 of 8 primary care clinics (vs secondary care clinic) with adjusted risk ratios (aRR) ranging from 1.45 to 2.31, patients diagnosed ≥90 days before facility-based HIV care enrolment (aRR 1.38; 1.01–1.88) (vs diagnosed on the same day as HIV care enrolment), and pregnant women (aRR 1.37; 1.15–1.62) (vs non-pregnant women).

### The effect of same-day ART initiation

#### Descriptive analyses

Crude decomposed outcomes and censoring due to TFO are presented in Figure 2, and Figure 3. The crude cumulative hazard of remaining on effective first-line ART (not experiencing the composite unfavourable outcome) was lower for same-day ART (vs early ART) during the first 2 years after ART initiation but comparable at 3 years (see Supplementary Figure 1). For same-day ART, it was 72% (95% CI: 68–74%) (vs early ART: 81%; 77–84%) at 1 year and 62% (59–66%) (vs early ART: 69%; 63–73%) at 3 years. The likelihood of experiencing the unfavourable outcome was high immediately after ART initiation, with 3.7% (95% CI: 2.3–5.8%) and 8.7% (95% CI: 7.0–10.8%) of patients under early and same-day ART never returning to care.

**Figure 3:**
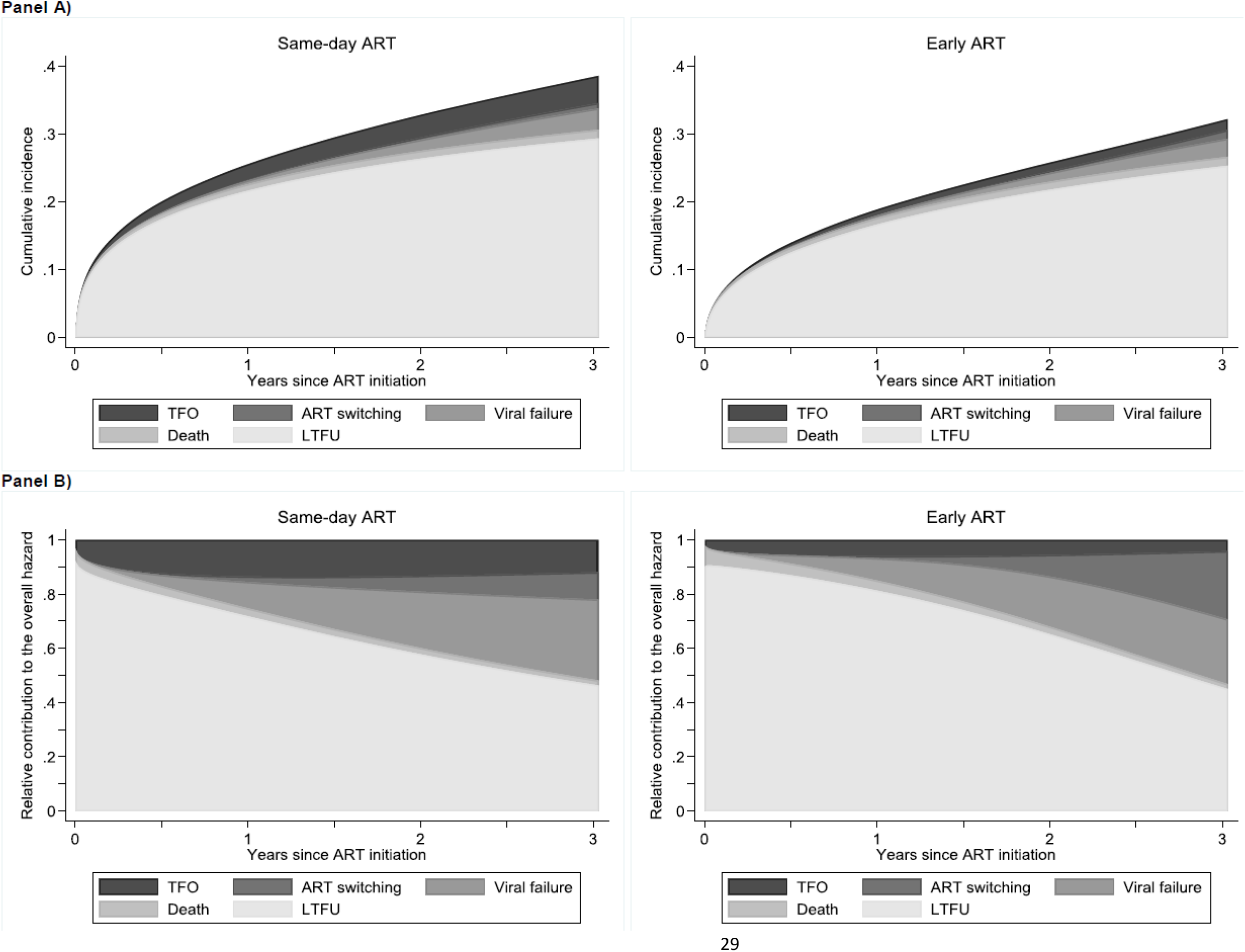
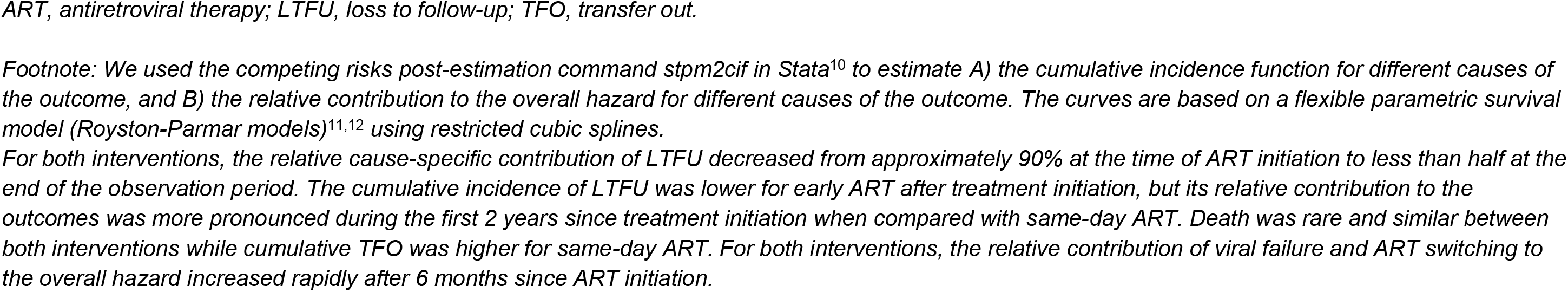
Stacked cause-specific cumulative incidence functions (Panel A), and stacked cause-specific relative contribution to the overall hazard (Panel B) of the outcomes of LTFU, death, viral failure, treatment switching and censoring due to TFO for early vs same-day ART, Same-day antiretroviral therapy under Treat-All, 2014–2016.

#### Relative impact of same-day ART on the unfavourable outcome

Multiple imputation of missing values was successful, with good convergence of the imputation algorithm and good other diagnostics (see Supplementary Figure 2 and 3). The hazard of the unfavourable treatment outcome was increased for same day-ART by 39% in univariate analysis (crude hazard ratio [cHR] 1.39; 95% CI: 1.14–1.70) and by 48% in multivariable analysis (adjusted hazard ratio [aHR] 1.48; 95% CI: 1.16–1.89) (Figure 4a) (see Supplementary Table 3 for the full model). The effect varied over time, with a higher hazard during the first year after ART initiation and a similar hazard thereafter (Figure 4b).

**Figure 4:**
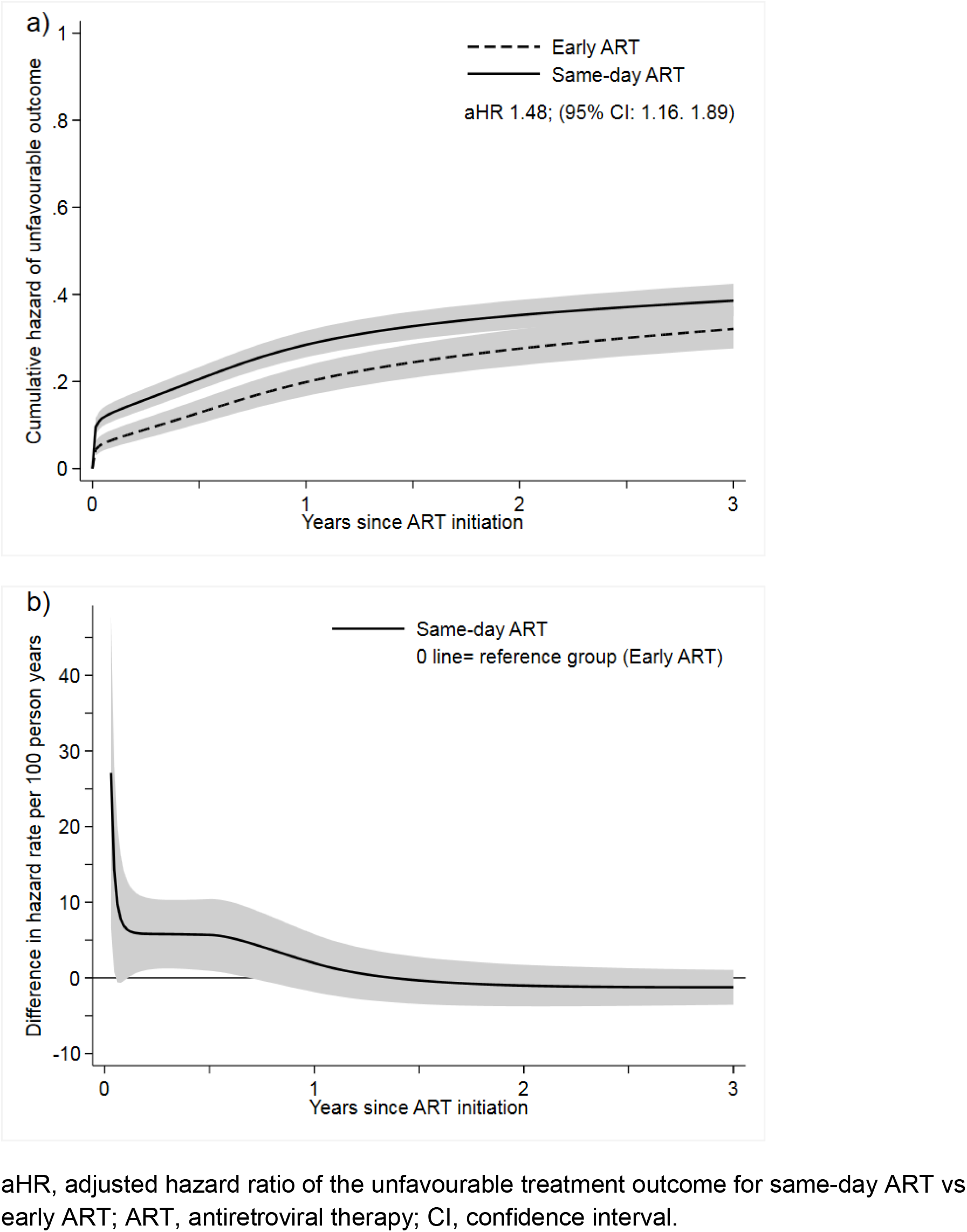
Averaged cumulative hazard (a) and averaged difference in hazard rate (b) of the unfavourable outcome for time from ART initiation to unfavourable outcome for patients initiating same-day ART vs early ART, Same-day antiretroviral therapy under Treat-All, 2014–2016.

#### Absolute difference in unfavourable outcomes comparing same-day ART with early ART

Using TMLE, we estimated that 28.9% (95% CI: 25.4–32.3%) of patients would have experienced an unfavourable outcome after 12 months if they had received same-day ART compared with 21.2% (15.8–26.6%) if they had received early ART, which corresponds to a difference of 7.7% (1.3–14.1%) and an risk ratio of 1.36 (1.03–1.81). Differences between the two treatment strategies were also observed for 2 and 3 years of follow-up, though less pronounced than in the first year (see Figure 5). Diagnostics of the TMLE approach were satisfactory, with no truncation of estimated probabilities of treatment assignment, small maximum clever covariates and a broad selection of learning algorithms (see Supplementary Table 2).

**Figure 5:**
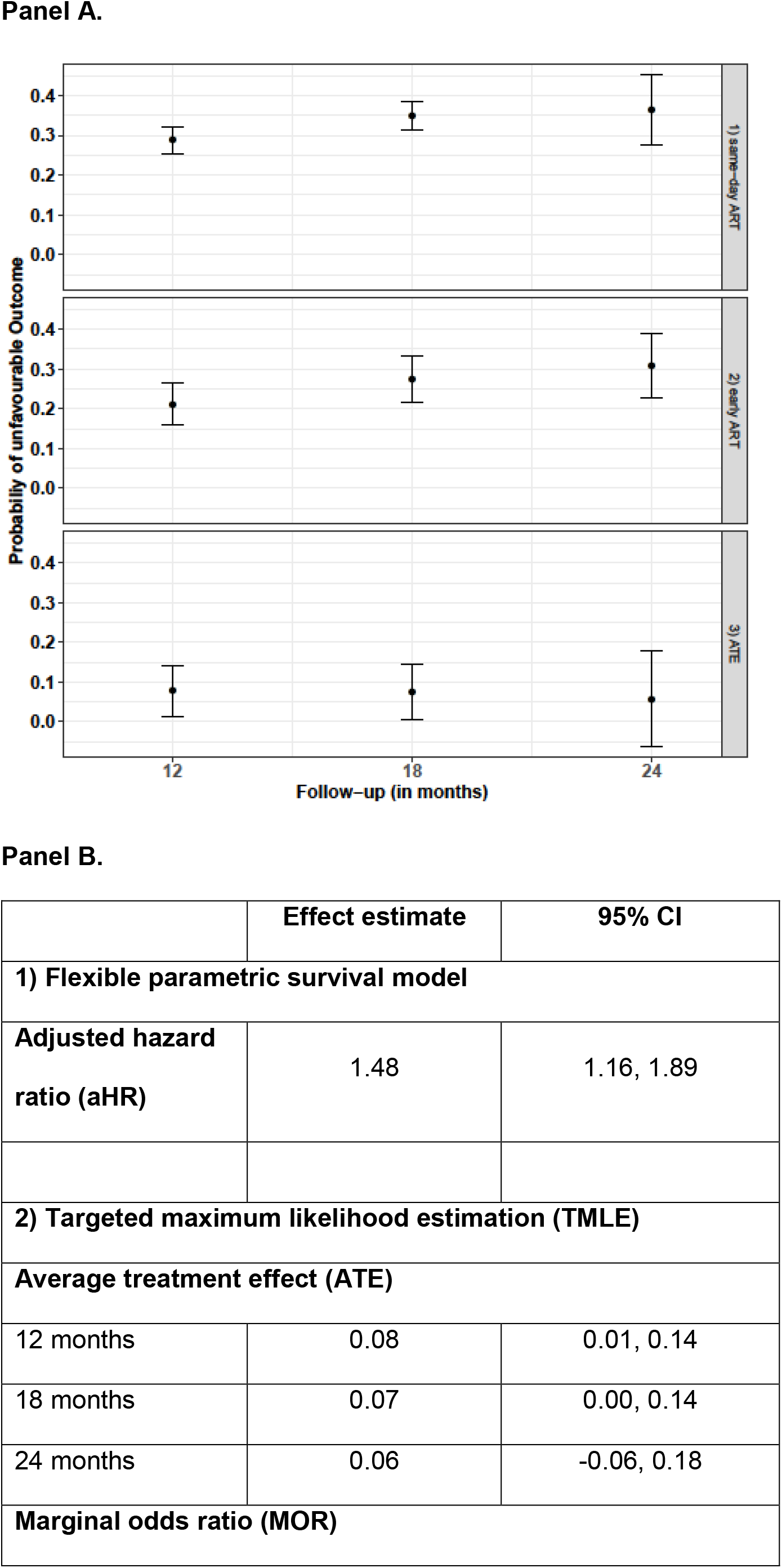
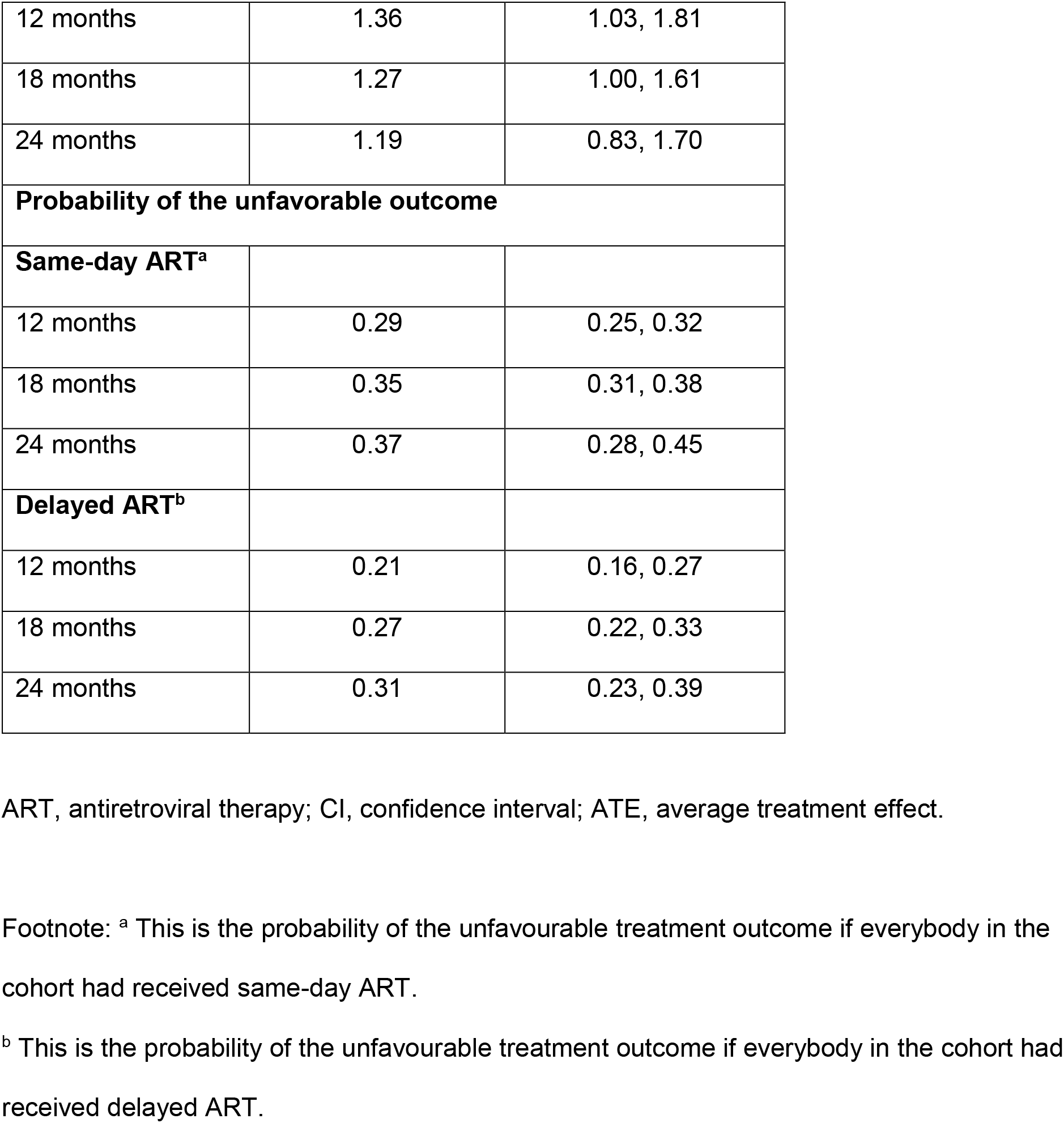
Main results: estimated effect of same-day ART initiation on the unfavourable outcome using TMLE (panels A and B) and using flexible parametric survival models (panel B), Same-day antiretroviral therapy under Treat-All, 2014–2016.

#### Supplementary analyses

Table 2 presents crude and adjusted hazard ratios for different assumptions. Changing time zero to the date of care enrolment, changing the unfavourable composite outcome to all-cause attrition and comparing same-day ART with rapid ART did not change findings overall, with adjusted hazard ratios ranging from 1.43 to 1.83.

**Table 2:**
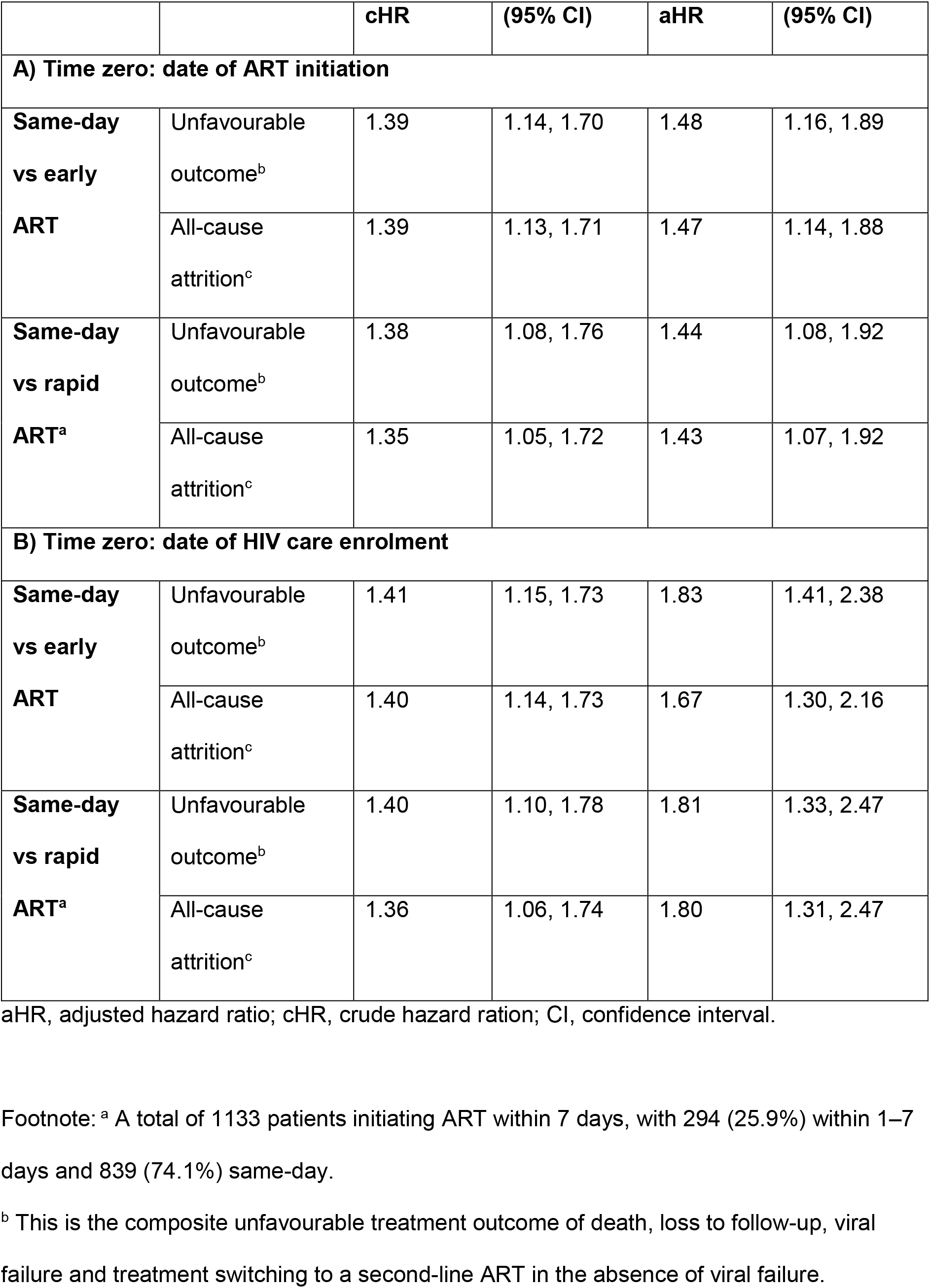
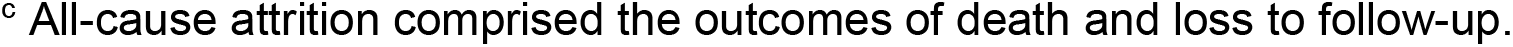
Effect estimates of combination of supplementary analyses with different assumptions, Same-day antiretroviral therapy under Treat-All, 2014–2016.

|  |  | **cHR** | **(95% CI)** | **aHR** | **(95% CI)** |
| --- | --- | --- | --- | --- | --- |
| **A) Time zero: date of ART initiation** |  |  |  |  |  |
| **Same-day vs early ART** | Unfavourable outcome<sup>b</sup> | 1.39 | 1.14, 1.70 | 1.48 | 1.16, 1.89 |
|  | All-cause attrition<sup>c</sup> | 1.39 | 1.13, 1.71 | 1.47 | 1.14, 1.88 |
| **Same-day vs rapid ART<sup>a</sup>** | Unfavourable outcome<sup>b</sup> | 1.38 | 1.08, 1.76 | 1.44 | 1.08, 1.92 |
|  | All-cause attrition<sup>c</sup> | 1.35 | 1.05, 1.72 | 1.43 | 1.07, 1.92 |
| **B) Time zero: date of HIV care enrolment** |  |  |  |  |  |
| **Same-day vs early ART** | Unfavourable outcome<sup>b</sup> | 1.41 | 1.15, 1.73 | 1.83 | 1.41, 2.38 |
|  | All-cause attrition<sup>c</sup> | 1.40 | 1.14, 1.73 | 1.67 | 1.30, 2.16 |
| **Same-day vs rapid ART<sup>a</sup>** | Unfavourable outcome<sup>b</sup> | 1.40 | 1.10, 1.78 | 1.81 | 1.33, 2.47 |
|  | All-cause attrition<sup>c</sup> | 1.36 | 1.06, 1.74 | 1.80 | 1.31, 2.47 |

aHR, adjusted hazard ratio; cHR, crude hazard ration; CI, confidence interval.
Footnote: <sup>a</sup> A total of 1133 patients initiating ART within 7 days, with 294 (25.9%) within 1–7 days and 839 (74.1%) same-day.
<sup>b</sup> This is the composite unfavourable treatment outcome of death, loss to follow-up, viral failure and treatment switching to a second-line ART in the absence of viral failure.
<sup>c</sup> All-cause attrition comprised the outcomes of death and loss to follow-up.

## DISCUSSION

This is to our knowledge the first study evaluating faster ART initiation in a routine programmatic HIV-care setting applying the Treat-All policy. In patients starting treatment quickly, initiating ART on the day of facility-based HIV care enrolment had inferior treatment outcomes compared with patients starting treatment 1–14 days thereafter or starting treatment within 1–7 days. The effect was accrued during the first year of therapy.

### Interpretation of findings

#### Predictors of same-day ART

The main predictors of same-day ART initiation were related to policy and facility factors. Pregnant women were associated with increased same-day ART initiation, coinciding with the same-day ART policy under PMTCTB+. Facility-level factors also played a role, with almost all primary care facilities providing more same-day ART than the secondary care facility. This may be because primary care facilities had point-of-care biochemistry, haemoglobin and CD4 testing available, thus making baseline results available on the same day for treatment decisions, as opposed to the secondary care facility where results often became available a few days later. Clinicians may have felt more comfortable initiating ART with CD4 cell count and biochemistry known. In addition, the one-stop-shop primary care clinics provided all HIV services at the same location whereas HIV testing and care registration were co-located in the secondary care facility. This required patients diagnosed with HIV in the outpatient department to transfer to the HIV department, thus possibly delaying care registration and ART initiation. More patients may also have had unmeasured co-morbidities at the secondary care facility, necessitating delaying ART initiation.

Patients who knew their HIV-positive status for ≥90 days were more likely to initiate ART on the same day. Firstly, patients may have been transferred in from community HIV testing sites and other facilities. Given more time between testing and care enrolment, they may have come to terms with life-long therapy and therefore been ready to start same-day treatment. Secondly, treatment interruptions are frequent in routine settings,^43^ and these patients may have been treatment interrupters re-initiating ART without disclosing prior treatment.

Lastly, clinical factors as well as social factors such as level of education and marital status appeared not to play a major role in quicker ART initiation. This may indicate that same-day ART initiation was driven by facility and health policy factors as indicated in our analysis rather than by clinical presentation of the patient, clinicians’ considerations, or patients’ preferences.

#### Effect of same-day ART

Same-day ART initiation had a higher hazard of the unfavourable treatment outcome than early and rapid ART. This effect was time-varying, with increased hazard during the first year of treatment and similar hazards thereafter.

We provide several explanations. Firstly, same-day ART may not address patients’ concerns about expedited ART initiation, and not give enough time to conceptualize lifelong therapy.^44–49^ This may have contributed to immediate disengagement from care after treatment initiation, with 9% of patients under same-day ART never returning for a follow-up visit.

Secondly, estimates may be affected by unmeasured confounding. Treatment readiness may predict assignment to the intervention and is also likely associated with the outcome (through the factor of adherence). In addition, we could not measure all possible baseline and time-updated co-morbidities that may predict the intervention and the outcome. For instance, cryptococcal meningitis may have been unevenly distributed in the groups and affect early death and loss to care differently.

Thirdly, the clinical tools used to assess treatment readiness may have been inappropriate to identify patients ready for same-day ART, as the very same tools were used before same-day ART initiation was an option. Contextualized screening tools for expedited ART initiation adapted to different populations (e.g. pregnant women) and settings may be needed to reliably assess patients’ readiness for same-day treatment. For instance, one randomized trial used a treatment readiness survey to identify patients not ready for same-day ART initiation and excluded them from expedited treatment.^16^ In addition, training related to expedited counselling protocols and same-day ART for health workers during the early implementation period was lacking, possibly leaving health workers poorly equipped for effective implementation of same-day ART at scale. Lastly, counselling support after sameday ART initiation may have been de-prioritized in this busy public sector setting with competing activities, thus providing insufficient adherence support early during treatment.

### Findings in context

The definition of same-day ART differs across studies. Definitions include treatment initiation on the day of HIV diagnosis, day of treatment eligibility, day of HIV care enrolment, or a combination of them.^15–17,50–52^ The same-day ART intervention group often consisted of patients initiating treatment days after the offer of same-day treatment,^15,17,51^ so that studies evaluated the intention to initiate same-day treatment rather than actual same-day treatment initiation.^15–17^ The offer of same-day ART was often combined with additional interventions (e.g. point-of-care CD4 and biochemistry testing),^53^ and restriction of the patient sample to healthier individuals^16^ and non-pregnant adults^15–17^ may make findings less applicable to routine public sector settings. Streamlining definitions of same-day ART initiation and clarity of what and who is evaluated are warranted.

While same-day ART initiation improves treatment uptake, it may downshift loss to care to the time of treatment.^15,53^ Treatment interruptions were already common in routine HIV programmes before the introduction of the rapid ART policy,^35,43^ and are associated with acquired drug resistance.^54^ Balancing of patient-level and public health benefits and risks (e.g. unstructured treatment interruptions) is required to make an informed health policy decision.

More emphasis may be needed on a differentiated approach to ART initiation adapted to the patient’s needs, with clinical and programmatic (e.g. logistical) constraints taken into consideration, than on choosing between same-day and rapid/early ART initiation as a blanket approach. In fragile health systems, hasty low quality and possibly coerced ART initiation may occur if HIV programmes and funding organizations prioritize achieving targets related to numbers of same-day ART initiations instead of differentiated patient-centred rapid ART initiation.

Importantly, this study did not assess the impact of a policy of same-day ART initiation for all PLHIV, as this was not feasible in our context (e.g. patients transferred in could not be offered same-day treatment), with the observational study design and available data. Thus, findings are not directly comparable to randomized trials evaluating the offer of same-day treatment to treatment-eligible patients. Our research, however, intends to estimate the risks and benefits of same-day ART initiation for patients with the ability to start treatment early. If there is a causal relationship between same-day ART and unfavourable treatment outcome, then deferral of treatment initiation should be considered for these patients. However, more research into the methods may be required to address questions of frequency, intensity, content and minimum quality of early adherence support in routine public sector settings.

### Limitations and strengths

Firstly, this study assessed outcomes of patients successfully initiated on ART soon after facility-based HIV care enrolment. Restriction allowed the establishment of two potentially comparable groups in the context of an observational study design but limits direct comparison with settings where most patients initiate ART 2 weeks after care enrolment. It was beyond the scope of this study to assess outcomes of patients starting treatment late or never, and they may differ in their characteristics and risks for an adverse outcome. By focusing only on one aspect of faster ART initiation, this study did not address the programmatic advantage of same-day ART in reaching patients otherwise defaulting before treatment. Future studies from the public sector should weigh the benefit of less pre-treatment loss with the risk of higher loss early during treatment.

Secondly, we did not account for loss between the diagnosis of HIV and care enrolment. This may have caused selection bias because only patients successfully linked to facility-based HIV care are considered. Specifically, loss between community-based HIV diagnosis and facility-based enrolment can be high,^17,55^ ranging from 10% to more than half in Eswatini.^12,50^ Intra-facility linkage in this setting may also be sub-optimal as estimated to be between 83% and 92%.^12,50,56^ Therefore, findings should not be generalized to predominantly community settings but rather to settings similar to ours where most HIV diagnosis happens at facility-level.^57^

Thirdly, patients under same-day ART never returning for refills after treatment initiation could have been silent patient-initiated (undocumented) transfers. The proportion of silent transfers ranges from 5% to 54% in patients documented as LTFU in Africa and is more pronounced in recent and larger treatment cohorts.^58^ We did not adjust for it because of a weak physical defaulter tracing intervention in place, and the inability of linking patient records to facilities outside the intervention area. Understanding the magnitude of silent transfer under Treat-All and if it differs between same-day and early ART should be further explored to inform health policy.

Fourthly, we could not adjust for all possible confounding factors identified in the DAG (e.g. co-morbidities and treatment readiness), possibly biasing the effect estimate in either direction.

A strength of this study is that we applied different analytical approaches, including state-of-the-art methods (TMLE), all of which concurred in their main findings. We included a wide range of patients as found in other HIV programmes implementing the Treat-All programmatic approach, so findings may be generalizable to similar settings in rural Sub-Saharan Africa. This study discussed potential shortfalls in programmatic implementation of Treat-All related to contextualized screening tools and training provided, thus drawing attention to the method and quality of implementation.

## Conclusions

Facility and health policy factors were the main predictors of same-day ART initiation. Our data also suggest that same-day ART increased the risk of the composite unfavourable outcome including LTFU. However, LTFU may sometimes relate to silent TFO, thus further research about true health outcomes of patients documented as lost to care is urgently needed.

## Data Availability

The software code and data can be obtained from the authors. New analyses on the data require approval from the government of Eswatini and MSF.

## Acknowledgements

We thank all the patients and healthcare workers who were involved in piloting the Treat-All approach and accelerated ART initiation in the Shiselweni region, and specifically the patients in Nhlangano health zone. In addition, we thank all the MSF teams involved in data collection and data cleaning.

## Supplementary Material

### Supplementary Technical appendix 1: Some explanation of the developed directed acyclic graph (DAG)

Treatment assignment, such as the decision to initiate ART on the same day, was based on the following considerations. Firstly, ART initiation on the day of facility-based HIV care enrolment was policy for pregnant/lactating women and encouraged for other patients in the absence of (presumptive) opportunistic infections. Secondly, same-day ART was practised at the clinician’s discretion based on the clinical and psychological pre-treatment assessment and depended on the patient’s self-perceived readiness, where the clinician’s advice may vary by facility. Thirdly, baseline CD4 cell count may also predict timing of ART initiation (e.g. patients with low CD4 count and no symptoms suggestive of opportunistic infection may initiate ART on the same day, patients with low CD4 cell count and symptoms suggestive of opportunistic infection may delay ART initiation and patients with high CD4 count may initiate immediately or delay). Fourthly, treatment readiness of the patient with regards to psychological preparedness (and not clinical readiness) at baseline could be a main factor predicting same-day ART initiation, as clinicians would not enforce same-day ART if the patient communicated not being ready for it. Fifthly, a patient’s marital status may also affect the decision to start treatment on the same day; married patients may favour deferring the decision on when to start treatment, as they may first wish to consult with their partner. However, higher education level may accelerate initiation, as knowledge of HIV and the benefit of treatment may be increased. Sixthly, the decision to start on the same day may relate to calendar year, as practising clinicians may change advice with increasing experience. Lastly, the time from HIV diagnosis to care enrolment may influence same-day ART, with patients knowing their HIV status for longer possibly being more ready to start treatment immediately. For these reasons, we considered sex, pregnancy status, age, co-morbidities at enrolment, laboratory values at enrolment, education, marital status, temporal trends (calendar year), time of HIV diagnosis, facility, treatment counselling and treatment readiness to be relevant to deciding on the timing of treatment assignment. Consequently, these variables lie on back-door paths from treatment to the outcome.^1^

There are two ways through which timing of ART initiation could affect the composite outcome. The first way is biologically, if treatment delay would affect viral suppression and thus the development of co-morbidities and negative outcomes. Secondly, earlier treatment may have a psychological impact on patients, who, if they did not feel ready for ART and were pushed into treatment, could stop being adherent to their treatment or even be lost to the programme.

Note that all post-treatment variables, which are visualized in the DAG, are mediators on the path from the intervention to the outcome and should thus not be conditioned upon.^1^ However, those variables that determine treatment assignment, as described above, are crucial to block back-door paths from the intervention to the outcome. While we have measured most of these variables, treatment readiness and counselling are unmeasured, as are some baseline comorbidities; this suggests the possibility of some unmeasured confounding.

**Supplementary Table S1:** Pre-treatment variables with missing values, Same-day antiretroviral therapy under Treat-All, 2014–2016.

| Variable | Complete observations | Incomplete observations |  |
| --- | --- | --- | --- |
|  |  | n | % |
| CD4 count | 1276 | 52 | 4.1 |
| Body-mass-index | 1207 | 121 | 10.0 |
| Haemoglobin | 1008 | 320 | 31.7 |
| ALT | 1029 | 299 | 29.1 |
| Creatinine | 1096 | 232 | 21.2 |
| Time from HIV diagnosis to care enrolment | 1320 | 8 | 0.6 |
| WHO clinical stage | 1317 | 11 | 0.8 |
| Education | 1116 | 212 | 19.0 |
| Pregnancy status | 1321 | 7 | 0.5 |
| Timing of HIV diagnosis | 1322 | 6 | 0.5 |
| Marital status | 1302 | 26 | 2.0 |
| Phone availability | 1311 | 17 | 1.3 |

*ALT, alanine transaminase; n, number; %, percentage.*

**Supplementary Table S2:** Details and diagnostics for the TMLE analysis, Same-day antiretroviral therapy under Treat-All, 2014–2016. The diagnostics were based on a TMLE analysis using the R-package *ltmle*,^2^ with 10-fold cross validation, a squared loss function (for cross validation), and a truncation level of 0.01, meaning that estimated treatment assignment and censoring probabilities (needed for the targeted update step) would have been truncated if they were lower than 0.01 (although this did not occur). Results averaged over the 10 imputed datasets are reported.

|  | 12 months |  | 24 months |  | 36 months |  |
| --- | --- | --- | --- | --- | --- | --- |
|  | Same day | Early | Same day | Early | Same day | Early |
| Number uncensored and followed treatment | 814 | 483 | 810 | 482 | 570 | 395 |
| % truncated | 0% | 0% | 0% | 0% | 0% | 0% |
| Mean CC | 0.89 | 0.75 | 0.89 | 0.75 | 0.72 | 0.66 |
| Max. CC | 3.86 | 9.76 | 4.09 | 12.25 | 4.91 | 8.69 |

|  |  | 12 months |  |  | 24 months |  |  | 36 months |  |  |
| --- | --- | --- | --- | --- | --- | --- | --- | --- | --- | --- |
|  | Model | Outc. | Treat. | Cens. | Outc. | Treat. | Cens. | Outc. | Treat. | Cens. |
| Learner | Screening |  |  |  |  |  |  |  |  |  |
| SL.mean | -- | 0.215 | 0.000 | 0.000 | 0.252 | 0.000 | 0.000 | 0.076 | 0.000 | 0.000 |
| SL.glm | -- | 0.051 | 0.216 | 0.000 | 0.000 | 0.212 | 0.000 | 0.020 | 0.205 | 0.283 |
| SL.bayesglm | -- | 0.113 | 0.000 | 0.000 | 0.080 | 0.000 | 0.000 | 0.265 | 0.000 | 0.003 |
| SL.stepAIC | -- | 0.061 | 0.509 | 0.021 | 0.078 | 0.484 | 0.019 | 0.072 | 0.504 | 0.031 |
| SL.gam | -- | 0.000 | 0.000 | 0.002 | 0.000 | 0.000 | 0.019 | 0.002 | 0.000 | 0.072 |
| SL.knn | -- |  | 0.000 | 0.075 |  | 0.000 | 0.075 |  | 0.000 | 0.007 |
| SL.lae | -- | 0.000 | 0.000 | 0.000 | 0.012 | 0.000 | 0.026 | 0.028 | 0.000 | 0.000 |
| SL.randomForest | -- | 0.160 | 0.191 | 0.205 | 0.082 | 0.193 | 0.196 | 0.152 | 0.173 | 0.083 |
| SL.nnet | -- | 0.014 | 0.000 | 0.608 | 0.041 | 0.001 | 0.427 | 0.143 | 0.000 | 0.000 |
| SL.bayesglm | Cramer | 0.000 | 0.000 | 0.000 | 0.019 | 0.000 | 0.007 | 0.000 | 0.000 | 0.024 |
| SL.gam | Cramer | 0.000 | 0.000 | 0.000 | 0.014 | 0.000 | 0.005 | 0.029 | 0.000 | 0.225 |
| SL.step.interaction | Cramer | 0.033 | 0.000 | 0.009 | 0.000 | 0.000 | 0.000 | 0.029 | 0.000 | 0.000 |
| SL.bayesglm | Lasso | 0.000 | 0.000 | 0.000 | 0.000 | 0.000 | 0.075 | 0.028 | 0.000 | 0.000 |
| *SL.gam* | *Lasso* | 0.000 | 0.000 | 0.000 | 0.000 | 0.000 | 0.058 | 0.000 | 0.000 | 0.018 |
| *SL.step.interaction* | *Lasso* | 0.000 | 0.000 | 0.076 | 0.000 | 0.000 | 0.068 | 0.030 | 0.000 | 0.047 |
| *SL.bayesglm* | *Lasso2* | 0.200 | 0.000 | 0.000 | 0.261 | 0.000 | 0.000 | 0.000 | 0.000 | 0.000 |
| *SL.gam* | *Lasso2* | 0.109 | 0.000 | 0.000 | 0.138 | 0.000 | 0.000 | 0.000 | 0.021 | 0.087 |
| *SL.step.interaction* | *Lasso2* | 0.026 | 0.084 | 0.000 | 0.000 | 0.110 | 0.000 | 0.025 | 0.097 | 0.031 |
| *SL.earth* | *Cramer* | 0.016 | 0.000 | 0.000 | 0.019 | 0.000 | 0.017 | 0.008 | 0.000 | 0.042 |
| *SL.earth* | *Lasso* | 0.000 | 0.000 | 0.003 | 0.003 | 0.000 | 0.006 | 0.093 | 0.000 | 0.048 |

**Supplementary Table S3:** Univariate and multivariate analysis of time to unfavourable treatment outcome for patients initiating same-day ART vs early ART, Same-day antiretroviral therapy under Treat-All, 2014–2016.

| (% missing values) | Univariate analysis<br>(n=1328) |  | Multivariate analysis<br>(n=1328)<sup>1</sup> |  |
| --- | --- | --- | --- | --- |
|  | cHR | 95% CI | aHR | 95% CI |
| **Intervention, (0%)** |  |  |  |  |
| Early ART | 1 |  | 1 |  |
| Same-day ART | 1.39 | 1.14, 1.70 | 1.48 | 1.16, 1.89 |
| **Year, (0%)** |  |  |  |  |
| 2014 | 1 |  | 1 |  |
| 2015 | 1.10 | 0.84, 1.43 | 1.12 | 0.84, 1.48 |
| 2016 | 1.27 | 0.91, 1.77 | 1.21 | 0.84, 1.75 |
| **Timing of HIV diagnosis, (0.5%)** |  |  |  |  |
| Pre Treat-All | 1 |  | 1 |  |
| Treat-All | 1.09 | 0.83, 1.41 | 0.76 | 0.47, 1.22 |
| **Facility, (0%)** |  |  |  |  |
| SHC | 1 |  | 1 |  |
| PHC-1 | 1.25 | 0.88, 1.78 | 1.17 | 0.80, 1.72 |
| PHC-2 | 1.32 | 0.92, 1.91 | 1.37 | 0.93, 2.01 |
| PHC-3 | 0.65 | 0.38, 1.10 | 0.65 | 0.38, 1.13 |
| PHC-4 | 0.89 | 0.60, 1.33 | 0.80 | 0.53, 1.23 |
| PHC-5 | 0.68 | 0.41, 1.12 | 0.64 | 0.37, 1.09 |
| PHC-6 | 1.40 | 1.04, 1.89 | 1.41 | 1.01, 1.97 |
| PHC-7 | 1.21 | 0.92, 1.61 | 1.17 | 0.85, 1.61 |
| PHC-8 | 1.02 | 0.65, 1.60 | 1.04 | 0.65, 1.64 |
| **Time from HIV diagnosis to care enrolment, (0.5%)** |  |  |  |  |
| Same-day | 1 |  | 1 |  |
| 1–89 days | 0.92 | 0.75, 1.14 | 1.00 | 0.80, 1.26 |
| ≥90 days | 0.89 | 0.68, 1.17 | 0.73 | 0.46, 1.15 |
| **Sex/pregnancy, (0.6%)** |  |  |  |  |
| Men | 0.91 | 0.71, 1.16 | 0.92 | 0.70, 1.22 |
| Non-pregnant women | 1 |  | 1 |  |
| Pregnant women | 1.30 | 1.05, 1.62 | 1.20 | 0.94, 1.54 |
| **Age at ART initiation, years, (0%)** |  |  |  |  |
| 16 to 24 | 1.44 | 1.17, 1.77 | 1.30 | 1.02, 1.65 |
| 25 to 49 | 1 |  | 1 |  |
| ≥50 | 0.76 | 0.49, 1.18 | 0.88 | 0.56, 1.39 |
| **Marital status, (2.0%)** |  |  |  |  |
| Married | 1 |  | 1 |  |
| Not married | 1.40 | 1.13, 1.74 | 1.28 | 1.02, 1.61 |
| **Education, (16.0%)** |  |  |  |  |
| None | 1 |  | 1 |  |
| Primary | 0.89 | 0.51, 1.56 | 0.91 | 0.51, 1.63 |
| Secondary | 1.09 | 0.64, 1.86 | 0.99 | 0.55, 1.78 |
| Tertiary | 1.04 | 0.41, 2.61 | 0.91 | 0.35, 2.40 |
| **CD4 count, cells/mm<sup>3</sup>, (3.9%)** |  |  |  |  |
| 0 to 100 | 1.31 | 0.97, 1.78 | 1.33 | 0.95, 1.86 |
| 101 to 200 | 0.95 | 0.68, 1.32 | 0.97 | 0.69, 1.37 |
| 201 to 350 | 1 |  | 1 |  |
| 351 to 500 | 1.26 | 0.94, 1.69 | 1.26 | 0.94, 1.69 |
| ≥501 | 1.25 | 0.93, 1.67 | 1.25 | 0.92, 1.68 |
| **WHO clinical stage, (0.8%)** |  |  |  |  |
| I/II | 1 |  | 1 |  |
| III | 0.85 | 0.65, 1.10 | 1.02 | 0.77, 1.36 |
| IV | 1.22 | 0.93, 1.60 | 1.54 | 1.09, 2.18 |
| **Tuberculosis, (0%)** |  |  |  |  |
| No | 1 |  | 1 |  |
| Yes | 0.78 | 0.50, 1.23 | 0.75 | 0.46, 1.21 |
| **BMI, kg/m<sup>2</sup>, (8.8%)** |  |  |  |  |
| <18.5 | 1.29 | 0.87, 1.93 | 1.23 | 0.81, 1.88 |
| 18.5 to 24.9 | 1 |  | 1 |  |
| ≥25 | 0.93 | 0.75, 1.16 | 0.90 | 0.71, 1.15 |
| **Haemoglobin, g/dL, (24.1%)** |  |  |  |  |
| ≤9 | 1.35 | 1.05, 1.73 | 1.21 | 0.93, 1.58 |
| ≥10 | 1 |  | 1 |  |
| **ALT, U/L, (22.5%)** |  |  |  |  |
| ≤42 | 1 |  | 1 |  |
| ≥43 | 0.88 | 0.62, 1.24 | 0.91 | 0.63, 1.30 |
| **Creatinine, μmol/L, 17.5%)** |  |  |  |  |
| ≤120 | 1 |  | 1 |  |
| ≥121 | 1.74 | 1.02, 2.98 | 1.96 | 1.13, 3.38 |
| **Phone availability, (1.3%)** |  |  |  |  |
| No | 1 |  | 1 |  |
| Yes | 0.84 | 0.61, 1.14 | 0.73 | 0.52, 1.02 |

ALT, alanine transaminase; aHR, adjusted hazard ratio; cHR, crude hazard ratio; ART, antiretroviral therapy; BMI, body mass index; n, number; PHC, primary health care facility; SHC, secondary health care facility; WHO, World Health Organization.
*Footnote: A standard first-line treatment regimen contained 3TC, TDF or AZT, and EFV or NVP. A baseline TB case was any incident TB case between 6 months before and 1.5 months after ART initiation. Baseline clinical and laboratory data were obtained at the time of ART initiation and categorized into normal and pathological. The covariate HIV diagnosis describes whether HIV-positive diagnosis was established before or during the roll-out of the Treat-All policy.*
<sup>1</sup>Model specifications: The flexible parametric survival model had 3 internal knots (4 degrees of freedom) and 1 internal knot (2 degrees of freedom) for the time-varying covariate intervention exposure.

**Supplementary Figure 1:**
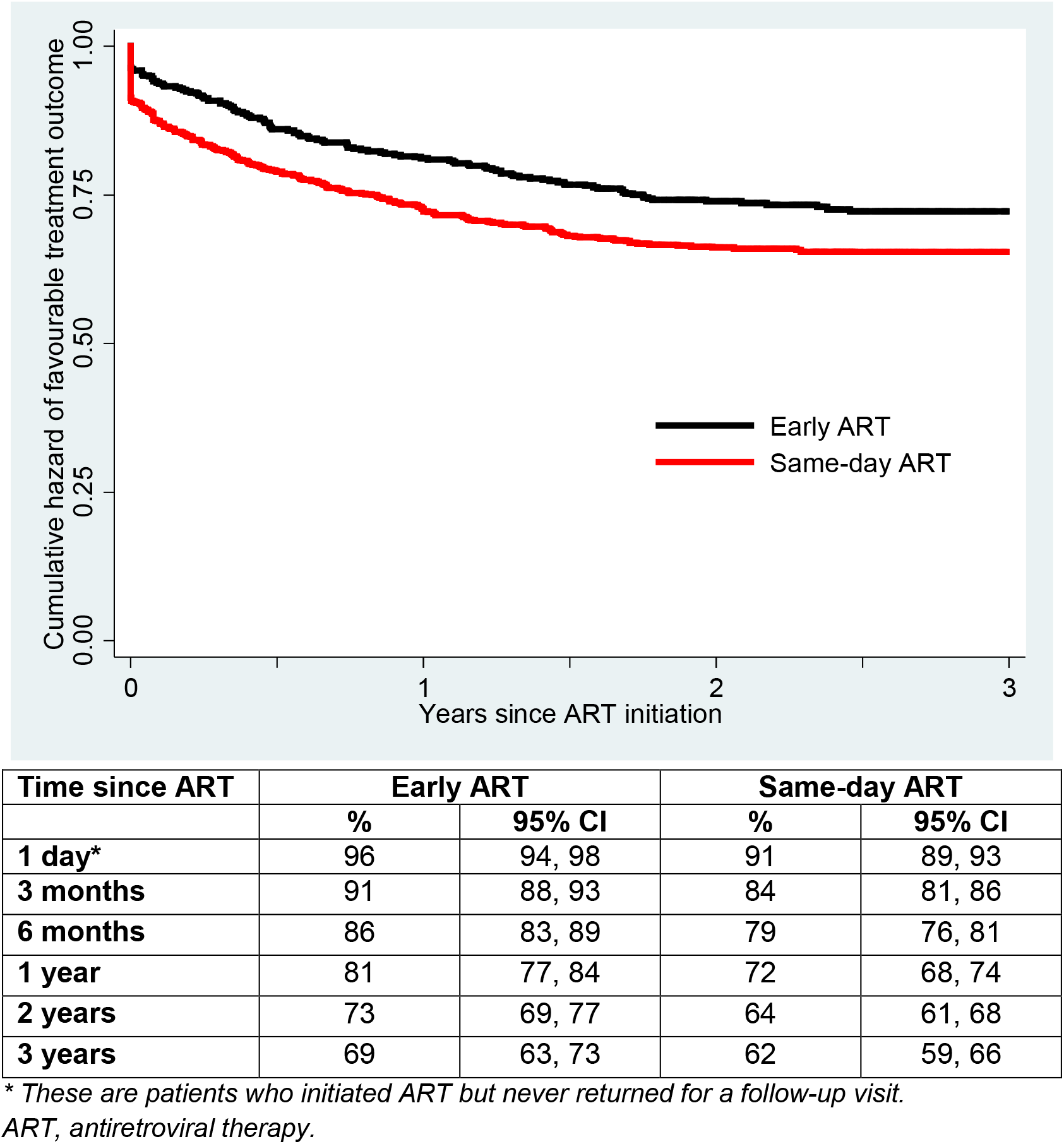
Cumulative hazard of favourable treatment outcome for patients initiating same-day ART vs early ART, Same-day antiretroviral therapy under Treat-All, 2014–2016.

**Supplementary Figure 2:**
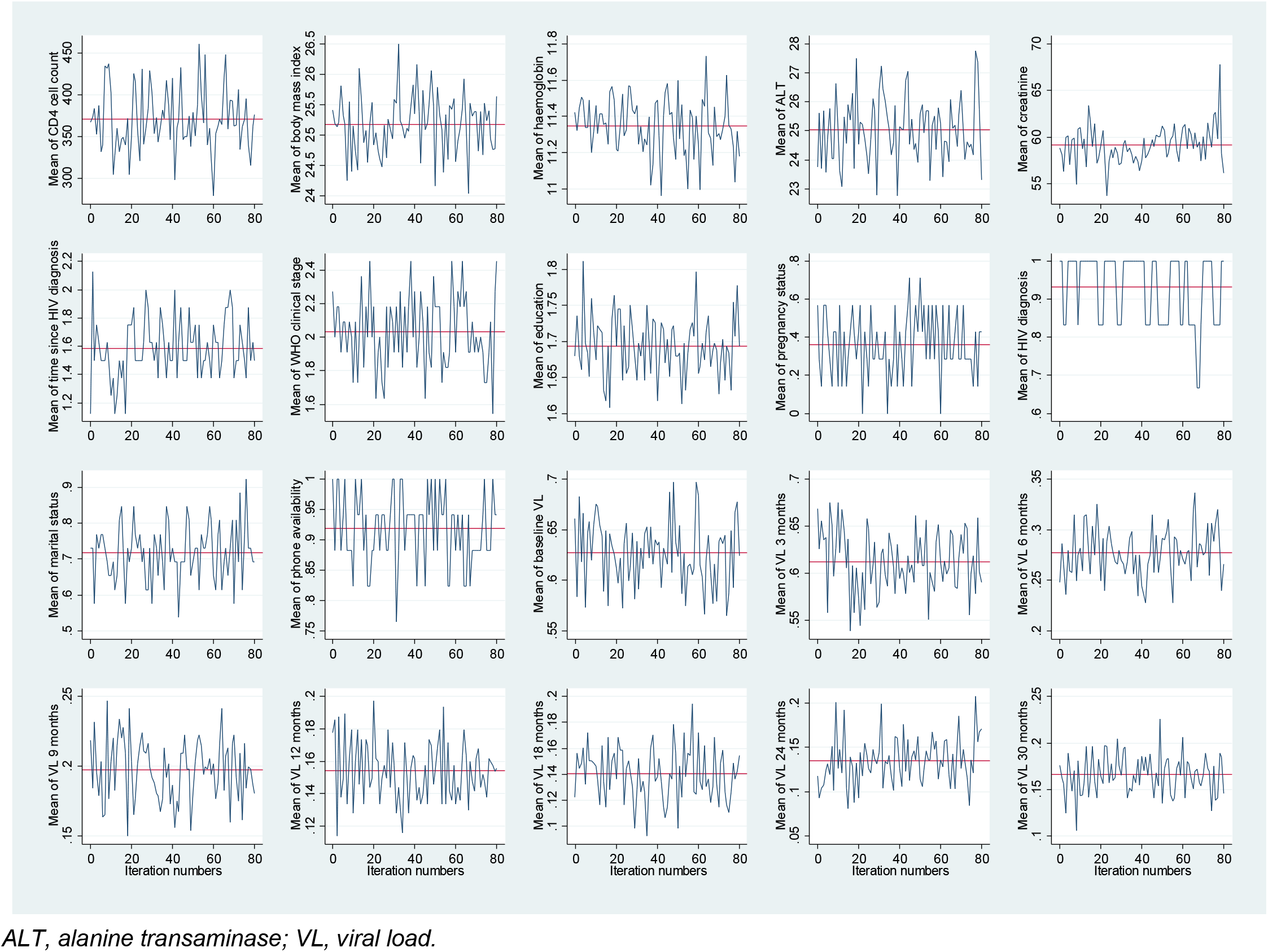
Trace plots of imputed data for all covariates with missing values, Same-day antiretroviral therapy under Treat-All, 2014–2016.

**Supplementary Figure 3:**
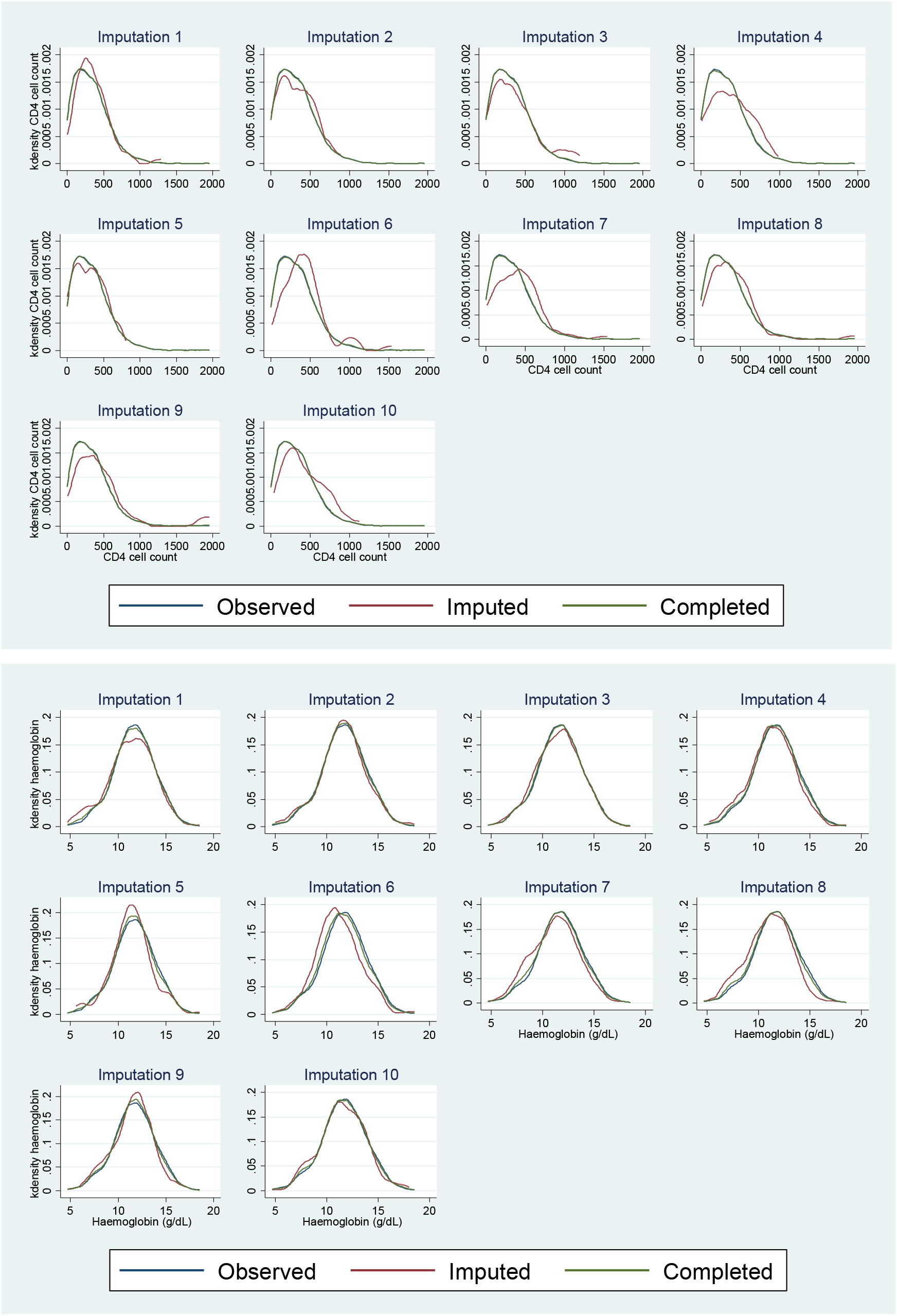
Kernel density plots for imputed CD4 cell count and haemoglobin for all imputed datasets as an example using the *midiagplots* command in Stata, Same-day antiretroviral therapy under Treat-All, 2014–2016.

